# Ethnic differences in COVID-19 mortality during the first two waves of the Coronavirus Pandemic: a nationwide cohort study of 29 million adults in England

**DOI:** 10.1101/2021.02.03.21251004

**Authors:** Vahé Nafilyan, Nazrul Islam, Rohini Mathur, Dan Ayoubkhani, Amitava Banerjee, Myer Glickman, Ben Humberstone, Ian Diamond, Kamlesh Khunti

## Abstract

**Background:** Ethnic minorities have experienced disproportionate COVID-19 mortality rates in the UK and many other countries. We compared the differences in the risk of COVID-19 related death between ethnic groups in the first and second waves the of COVID-19 pandemic in England. We also investigated whether the factors explaining differences in COVID-19 death between ethnic groups changed between the two waves.

**Methods:** Using data from the Office for National Statistics Public Health Data Asset on individuals aged 30-100 years living in private households, we conducted an observational cohort study to examine differences in the risk of death involving COVID-19 between ethnic groups in the first wave (from 24^th^ January 2020 until 31^st^ August 2020) and second wave (from 1^st^ September to 28^th^ December 2020). We estimated age-standardised mortality rates (ASMR) in the two waves stratified by ethnic groups and sex. We also estimated hazard ratios (HRs) for ethnic-minority groups compared with the White British population, adjusted for geographical factors, socio-demographic characteristics, and pre-pandemic health conditions.

**Results:** The study population included over 28.9 million individuals aged 30-100 years living in private households. In the first wave, all ethnic minority groups had a higher risk of COVID-19 related death compared to the White British population. In the second wave, the risk of COVID-19 death remained elevated for people from Pakistani (ASMR: 339.9 [95% CI: 303.7 – 376.2] and 166.8 [141.7 – 191.9] deaths per 100,000 population in men and women) and Bangladeshi (318.7 [247.4 – 390.1] and 127.1 [91.1 – 171.3] in men and women)background but not for people from Black ethnic groups. Adjustment for geographical factors explained a large proportion of the differences in COVID-19 mortality in the first wave but not in the second wave. Despite an attenuation of the elevated risk of COVID-19 mortality after adjusting for sociodemographic characteristics and health status, the risk was substantially higher in people from Bangladeshi and Pakistani background in both the first and the second waves.

**Conclusion:** Between the first and second waves of the pandemic, the reduction in the difference in COVID-19 mortality between people from Black ethnic background and people from the White British group shows that ethnic inequalities in COVID-19 mortality can be addressed. The continued higher rate of mortality in people from Bangladeshi and Pakistani background is alarming and requires focused public health campaign and policy changes.

*VN and NI contributed equally to this paper

**Research in context:** *Evidence before this study:* A recent systematic review by Pan and colleagues demonstrated that people of ethnic minority background in the UK and the USA have been disproportionately affected by the Coronavirus (COVID-19) pandemic, compared to White populations. While several studies have investigated whether adjusting for socio-demographic and economic factors and medical history reduces the estimated difference in risk of mortality and hospitalisation, the reasons for the differences in the risk of experiencing harms from COVID-19 are still being explored during the course of the pandemic. Studies so far have analysed the ethnic differences in COVID-19 mortality in the first wave of the pandemic. The evidence on the temporal trend of ethnic inequalities in COVID-19 mortality, especially those from the second wave of the pandemic, is scarce.

*Added value of this study:* Using data from the Office for National Statistics (ONS) Public Health Data Asset on 29 million adults aged 30-100 years living in private households in England, we conducted an observational cohort study to examine the differences in the risk of death involving COVID-19 between ethnic groups in the first wave (from 24^th^ January 2020 until 31^st^ August 2020) and second wave (from 1^st^ September to 28^th^ December 2020). We find that in the first wave all ethnic minority groups were at elevated risk of COVID-19 related death compared to the White British population. In the second wave, the differences in the risk of COVID-19 related death attenuated for Black African and Black Caribbean groups, remained substantially higher in people from Bangladeshi background, and worsened in people from Pakistani background. We also find that some of the factors explaining these differences in mortality have changed in the two waves.

*Implications of all the available evidence:* The risk of COVID-19 mortality during the first wave of the pandemic was elevated in people from ethnic minority background. An appreciable reduction in the difference in COVID-19 mortality in the second wave of the pandemic between people from Black ethnic background and people from the White British group is reassuring, but the continued higher rate of mortality in people from Bangladeshi and Pakistani background is alarming and requires focused public health campaign and policy response. Focusing on treating underlying conditions, although important, may not be enough in reducing the inequalities in COVID-19 mortality. Focused public health policy as well as community mobilisation and participatory public health campaign involving community leaders may help reduce the existing and widening inequalities in COVID-19 mortality.

## Introduction

A recent systematic review of 50 studies have showed that people from ethnic minority background in the UK and other countries, particularly Black and South Asian groups, have been disproportionately affected by the Coronavirus (COVID-19) pandemic compared to people of White ethnic background [1] While several studies have investigated whether adjusting for socio-demographic and economic factors and medical history reduces the estimated difference in risk of mortality and hospitalisation [2, 3, 4], the reasons for the differences in the risk of experiencing harms from COVID-19 are still being explored during the course of the pandemic. Factors including structural racism [5, 6], social vulnerability [7, 8] social and material deprivation, [9] have widely been suggested as potential mechanisms for these reported inequalities.

In view of changes in policy, treatments and roll out of vaccination programmes, understanding the evolving nature of the COVID-19 epidemiology is crucial in helping shape the public health response to the coronavirus pandemic, especially in the context of emerging variants in some countries. [10] As emerging evidence suggest that the long-term consequences of COVID-19 may be severe, especially amongst people from ethnic minority groups [11], it is critical to monitor how ethnic inequalities throughout the course of the pandemic have evolved.

Using nationwide population-level data containing detailed socio-demographic characteristics and information on pre-pandemic health status, we compared the difference in risk of COVID-19 related death between ethnic groups in the two waves of the COVID-19 pandemic. We also investigated whether the factors explaining differences in COVID-19 death between ethnic groups changed between the two waves. To our knowledge, it is the first study to examine how the difference in the COVID-19 mortality between ethnic groups changed when adjusting for both detailed socio-demographic factors and pre-pandemic health at a whole population level.

## Methods

### Data

Using data from the Office of National Statistics (ONS) Public Health Data Asset on approximately 29 million adults aged 30-100 years living in private households in England, we conducted an observational cohort study to examine the differences in the risk of death involving COVID-19 between ethnic groups in the first wave (from 24^th^ January 2020 until 31^st^ August 2020) and second wave (from 1^st^ September to 28^th^ December 2020) of the pandemic. Since data on socio-demographic factors are very scarce the healthcare datasets, we obtained these data from the 2011 Census. The 2011 Census was linked to the General Practice Extraction Service (GPES) Data for pandemic planning and research which contains primary care records for all individuals living in England in November 2019. This dataset was further linked to mortality records, Hospital Episode Statistics, using the NHS number. To obtain NHS numbers for the 2011 Census, the 2011 Census was linked to the 2011-2013 NHS Patient Registers using deterministic and probabilistic matching, with an overall linkage rate of 94.6%. We excluded patients (approximately 12.4%) who did not have a valid NHS number or were not in the GPES dataset, and therefore were likely to have migrated out of the country. Most socio-demographic factors were drawn from the 2011 Census, and therefore may not represent people’s circumstances at the beginning of the pandemic. To limit measurement error, we restricted the sample to adults over the age of 30 to limit the measurement error.

### Outcomes

The outcome was COVID-19 related death (either in hospital or out of hospital), defined as confirmed or suspected COVID-19 death as identified by ICD-10 codes U07.1 or U07.2 mentioned on the death certificate anywhere on the death certificate. We analysed deaths in two time periods based on the death of occurrence: 24^th^ January 2020 to 31^st^ August 2020 (wave 1) and 1^st^ September 2020 to 28^th^ December 2020 (wave 2). We used 1^st^ September as a cut-off date because the number of COVID-19 related death reached its lowest point in the week commencing 31^st^ August 2020 [12].

### Exposure

The exposure of interest was self-reported ethnicity obtained from the 2011 Census. We used a 10-category classification [13] and used the White British ethnic group as the reference category in all models. Ethnicity was imputed in 3.0% of 2011 Census returns due to item non-response using nearest-neighbour donor imputation, the methodology employed by the Office for National Statistics across all 2011 Census variables.

### Covariates

Other covariates used in the regression models include socio-demographic characteristics (age, sex, index of multiple deprivation, housing, household composition, occupational exposure), geographical factors, and pre-pandemic health status (BMI, learning disability, cancer, and immunosuppression, and other health conditions). Geographical factors were based on the 2019 Patient Register; socio-demographic characteristics were obtained from the 2011 Census (since this is the most reliable source for these variables); BMI and comorbidities were derived based on the primary care and hospitalisation data and defined using the QCOVID risk prediction model [14]. Details of these variables are available in the Supplementary Table A1.

We hypothesised that each of these factors may be associated with the risk of COVID-19 mortality by either increasing the risk of becoming infected and/or the risk of mortality once infected with COVID-19.

### Statistical analyses

As a measure of differences in absolute risk of COVID-19 mortality, we calculated age-standardized mortality rates (ASMRs) for the different ethnic groups, whereby the age distribution within each group was standardized to the 2013 European Standardised Population. We calculated ASMRs separately for men and women.

The differences in the risk of COVID-19-related death across ethnic groups could be mediated by geographical factors, socio-demographic characteristics and pre-pandemic health. These factors fall on the causal path between ethnicity and COVID-19 mortality in a directed acyclic graph. To assess whether these factors accounted for some of the difference in risk between ethnic groups, we estimated Cox’s proportional hazards models adjusted for a range of factors. First, we estimated models that only adjusted for age. The age-adjusted hazard ratios (HRs) can be interpreted as a measure of inequality in COVID-19 mortality. We then added groups of control variables (geographical factors, socio-demographic characteristics, and pre-pandemic health) step by step and assessed how these affected the estimated HRs. When fitting the Cox models, we included all individuals who died during the analysis period and a weighted random sample of those who did not, with a sampling rate of 1% for those of white British ethnicity and 10% for adults from ethnic minority groups.

## Results

### Characteristics of the study population

Our analytical sample consisted of 28,946,702 people aged 30-100 years who were alive on 24 January 2020 and living in England in private households. The number of COVID-19 related deaths was 29,303 and 17,487 in the first (24^th^ January 2020 to 31^st^ August 2020) and second wave (1^st^ September 2020 to 28^th^ December 2020) of the pandemic, respectively.

In this cohort of people living in private households, 53% were women and the average age was 56 (SD: 16) years. 83% percent of individuals identified as people from the White British ethnic group. The gender and age distribution of those who had a COVID-19 related death was similar in the two periods. In the first period, women accounted for 40.8 per cent of COVID-19 related death, and the mean age at death was 79(12) years. In the second period, women accounted for 41% of COVID-19 related death and the mean age at death was 79 (11) years. The mean age at death remained similar in the two waves for all ethnic group (See Supplementary Table A2). A higher proportion of COVID-19 related death occurred amongst people from White British ethnic background in wave 2 (87.6%) compared to wave 1 (83.6%), while the proportion of death decreased from 1.4% in wave 1 to 0.4% in wave 2 among people from Black African ethnic group, and 2.4% to 0.9% among people from Black Caribbean ethnic background. The proportion of deaths increased with the level of index of multiple deprivation deciles (Table 1).

**Table 1.**
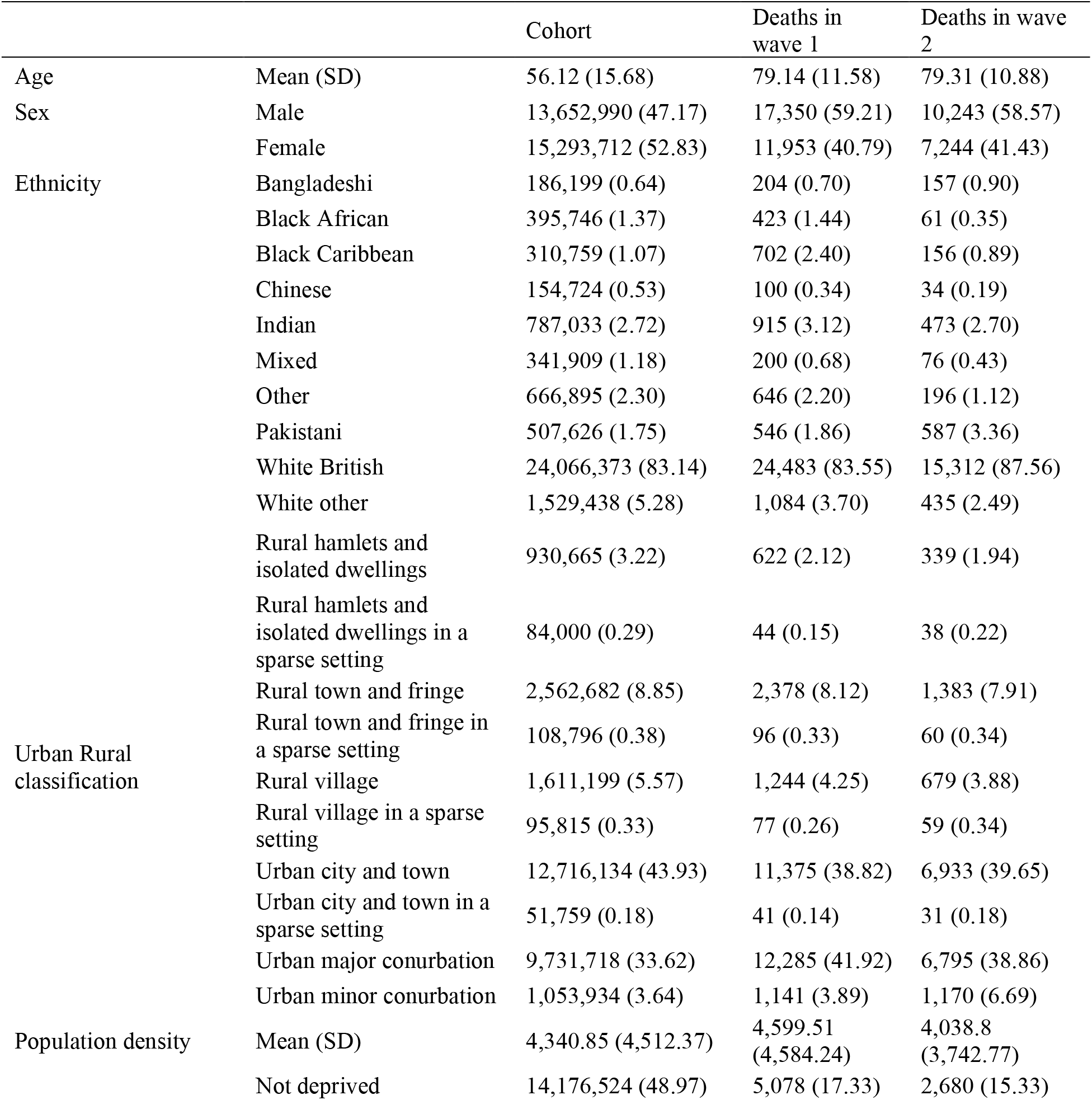
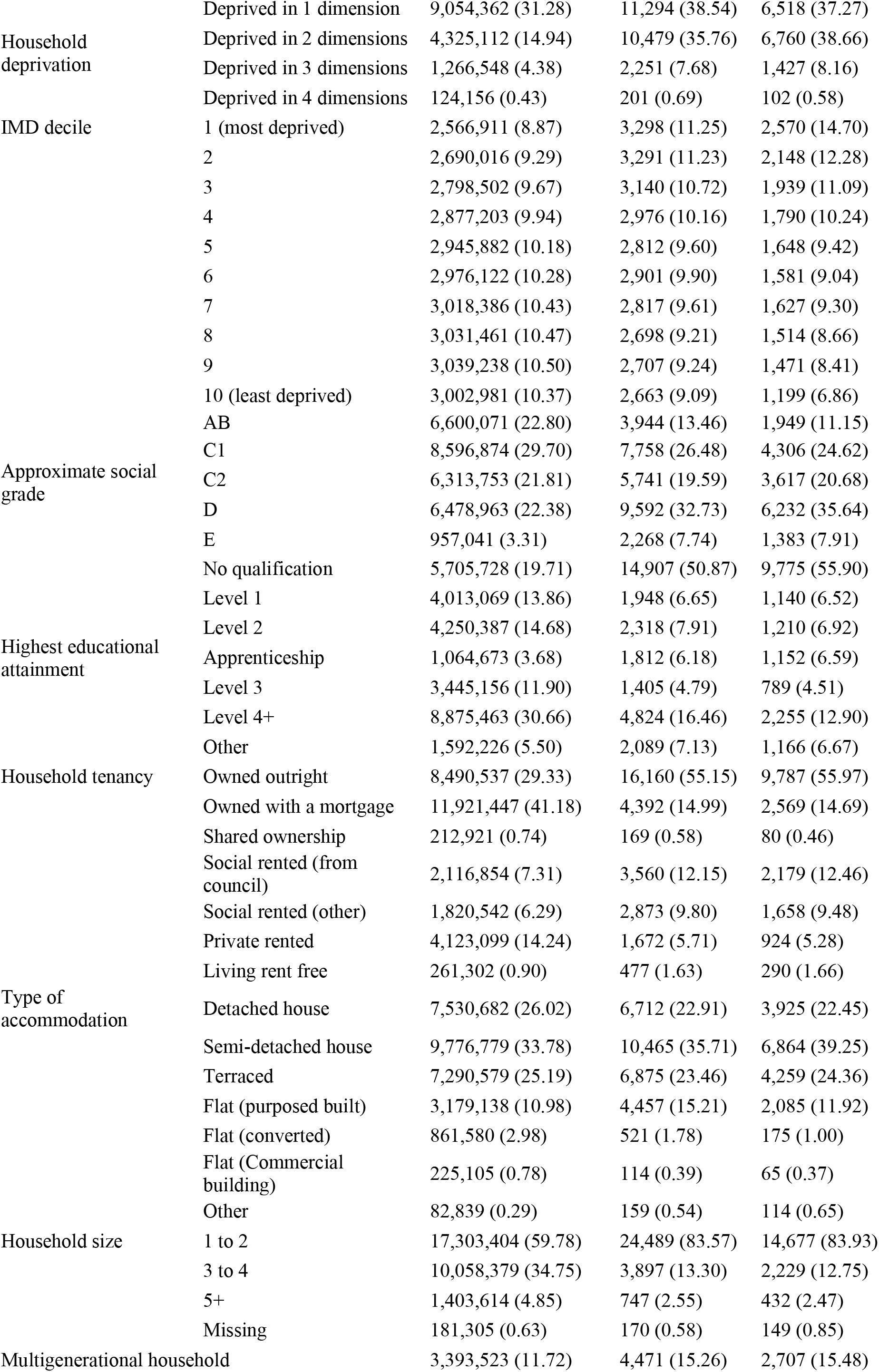
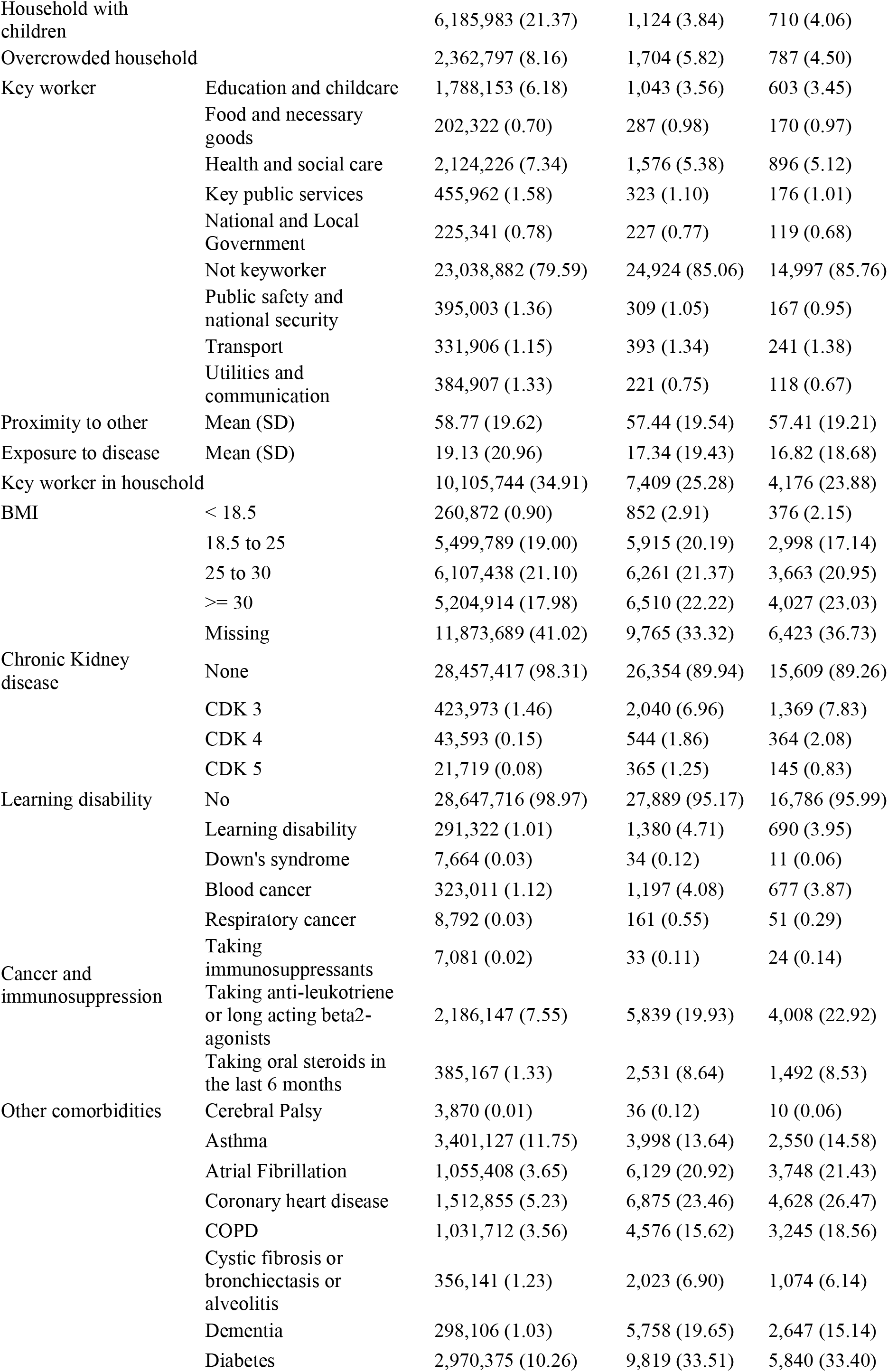
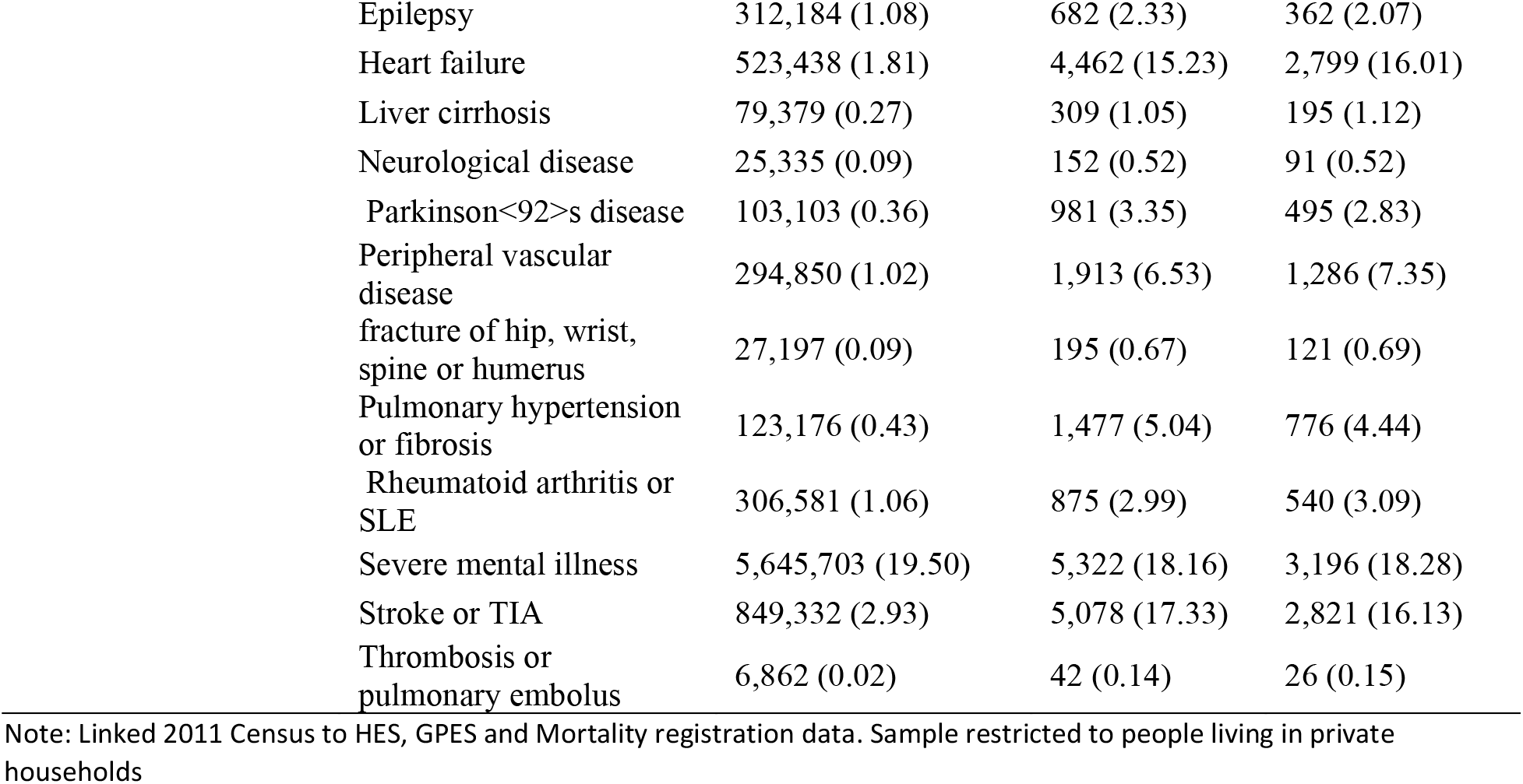
Demographic and medical characteristics for the study cohort and those who died with COVID-19 in the two waves

### Differences in COVID-19 mortality in wave 1 and wave 2: Age-standardized mortality rates

Table 2 shows the age-standardized mortality rates (ASMR) by ethnic group separately for the first and the second waves of the pandemic. In the first wave, the ASMRs of COVID-19 mortality were greatest among individuals identifying as Black African (402.5 [95% CI 341.6 – 463.4] and 174.4 [CI 137.6 – 210.5] deaths per 100,000 population in men and women, respectively). The ASMRs were lowest among those identifying as White British (119.1 [117.1 – 121.1] and 65.1 [63.8 – 66.3] deaths per 100,000 population in men and women, respectively). Levels of absolute risk were greater among all ethnic-minority groups compared with the White British population.

**Table 2.**
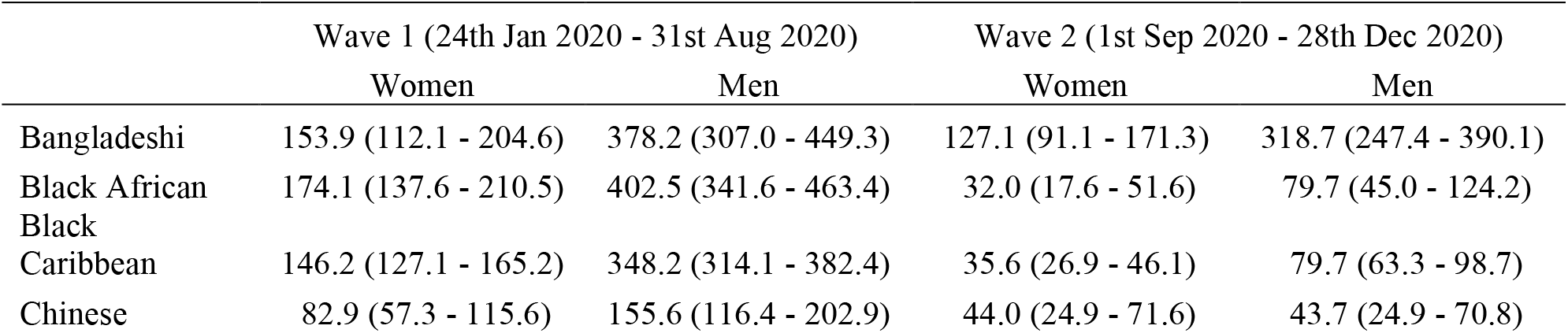
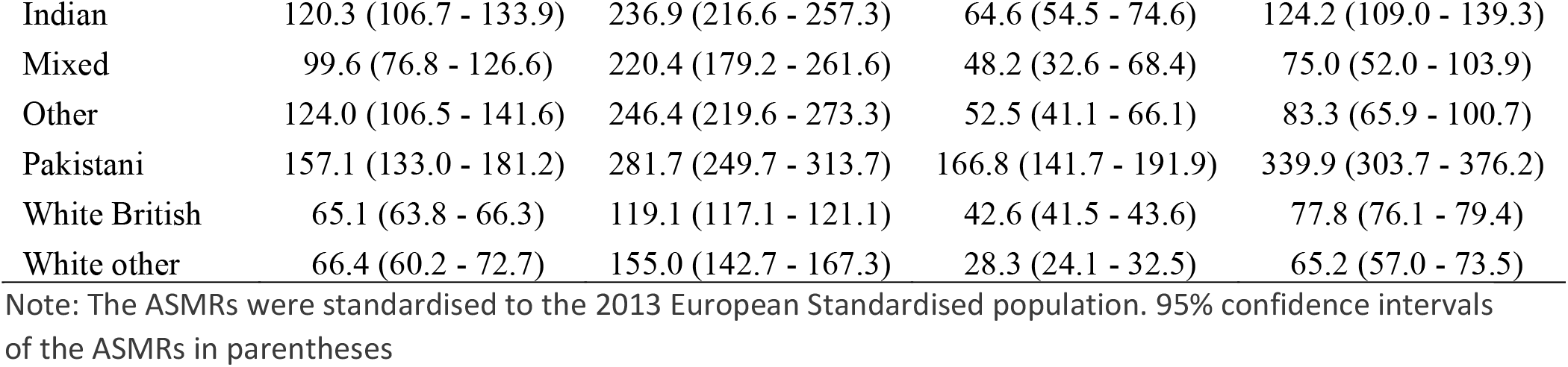
Age standardised mortality rates (ASMRs) of death involving COVID-19 per 100,000 population, stratified by sex and ethnic group

In the second wave, the ASMRs of COVID-19 mortality were highest among men and women identifying as Pakistani (339.9 [303.7 – 376.2] and 166.8 [141.7 – 191.9] deaths per 100,000 population in men and women) and Bangladeshi (318.7 [247.4 – 390.1] and 127.1 [91.1 – 171.3] deaths per 100,000 population in men and women) ethnic background. The ASMRs of COVID-19 mortality were lowest for people from other White background (65.2 [57.0 – 73.5] and 28.3 [24.1 – 32.5] deaths per 100,000 population in men and women) and the White British population (65.2 [57.0 – 73.5] and 28.3 [24.1 – 32.5] deaths per 100,000 population in men and women). Unlike in the first period, the ASMRs of COVID-19 mortality for people from Black African and Black Caribbean were similar to the ASMRs for people from the White British group.

### Determinants of disparities in COVID-19 mortality between ethnic groups

Figure 1 reports hazard ratios (HR) of COVID-19 related death in the first wave and second wave in men and women for ethnic minority groups compared with the White British population.

**Figure 1.**
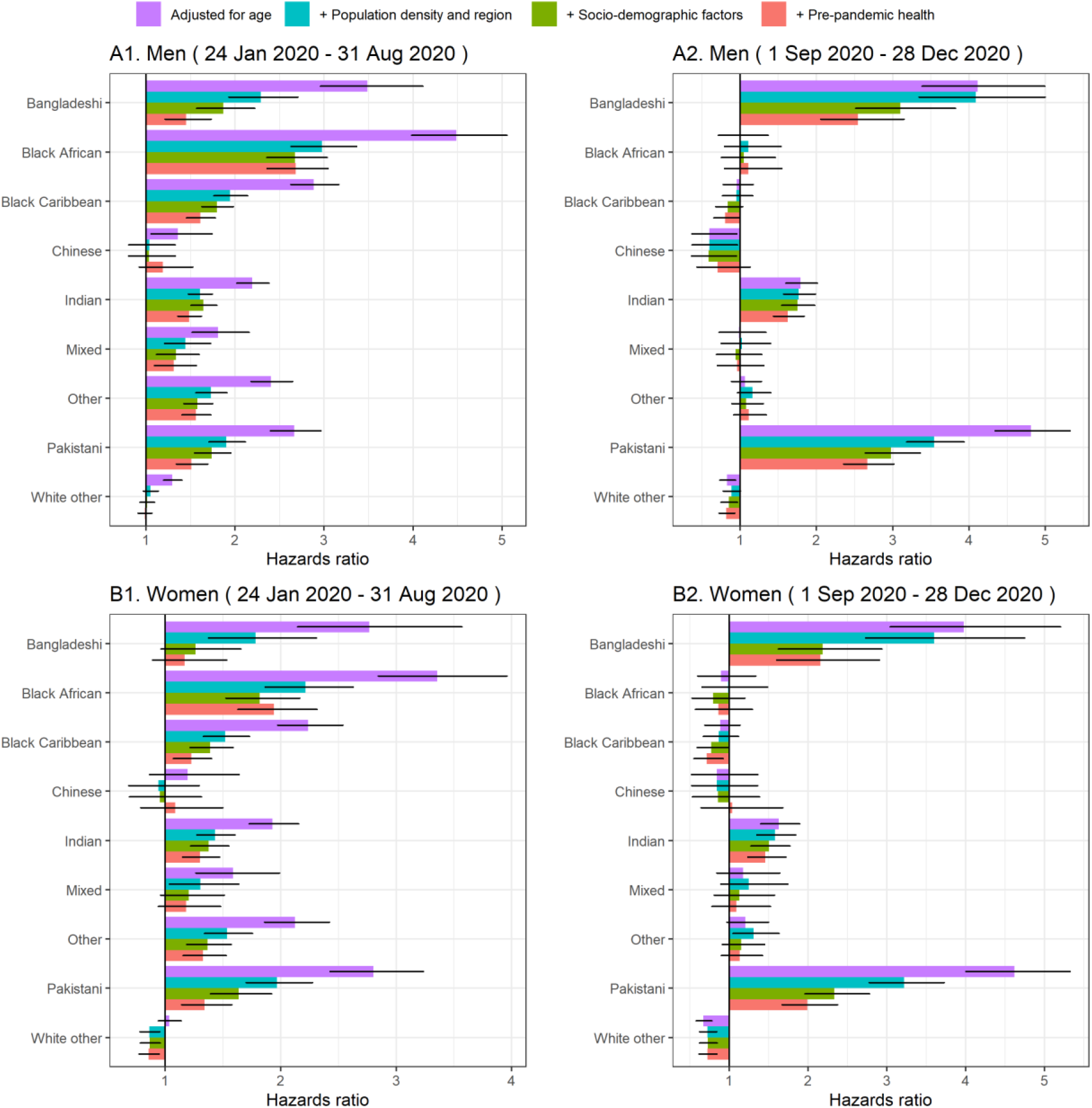
Hazard ratios for COVID-19 related death for ethnic-minority groups compared with the White British population, stratified by sex and pandemic waves. Note: Results obtained from Cox-regression models. Geographical factors: dummies for region of residence, for urban/rural classification and second order polynomial of population density of Lower Super Output Area (LSOA). Socio-demographic characteristics include Index of Multiple Deprivation (IMD), household deprivation (see table note), household tenure, social grade, level of highest qualification, household size, multigenerational household, household with children, key worker type, key worker in the household, exposure to disease, proximity to others, household exposure to disease, household proximity to others. Pre-pandemic health include Body Mass Index (kg/m2), Chronic kidney disease (CKD), Learning disability, Cancer and immunosuppression, other conditions (See Supplementary Tables A1 for more details). Numerical results can be found in Supplementary Tables A3)

As indicated by the ASMRs, age-adjusted HRs indicated that men and women from all ethnic-minority groups (except women of Chinese and White Other ethnicity) were at greater risk of COVID-19 related death compared with those of White British ethnicity in the first wave. The highest risk of mortality was observed among people from Black African ethnic background. For example, compared with men from White ethnic background, the rate of COVID-19 related deaths in wave 1 was 4.49 (95% confidence interval [CI]: 3.98-5.07) times higher in men from Black African ethnicity. In wave 2, men and women from South Asian ethnic groups were at greater risk of death involving COVID-19 compared with those of White British ethnicity (Figure 1), with adjusted HRs of 4.81 [4.34 – 5.32] and 4.62 [4.01 – 5.33] in men and women from Pakistani background, and 4.11 [3.38 - 4.99] and 3.98 [3.04 -5.20] in men and women from Bangladeshi background, respectively. Individuals from Indian background also had elevated risk of COVID-19 related death, with adjusted HRs of 1.80 [1.60 – 2.01] and 1.63 [1.40 – 1.90] in men and women, respectively. Unlike wave 1, people from Black ethnic groups were not at greater risk of COVID-19 death compared to those of White British ethnicity.

In both waves, adjusting for geographical factors, socio-demographic characteristics and pre-pandemic health substantially reduced the estimated disparities between most ethnic groups and the White British population. This suggests that the differences in mortality between ethnic groups are partly mediated by these factors. However, these factors attenuated the hazard ratios more strongly in Wave 1 than in wave 2. In addition, the factors that most strongly affected the HRs differed in the two waves.

In Wave 1, adjusting for geographical factors more than halved the estimated hazard ratios for all ethnic minority groups. For most groups, the hazard ratios were further reduced by adjusting for socio-demographic factors and pre-pandemic health status, especially amongst women. After adjusting for all these factors, women from Bangladeshi and Mixed background were no longer at greater risk of COVID-19 related death. For women from all other groups except Black African, the fully adjusted hazard ratios were below 1.4. However, despite the attenuation of the hazard ratios after full adjustment, men from all ethnic minority groups but other White remained at greater risk, but with hazard ratios greatly attenuated.

In Wave 2, adjusting for geographical factors did not substantially reduce the HRs in men and women from Bangladeshi background, but attenuated the HRs for people from Pakistani background. Adjusting for socio-demographic factors attenuated the elevated risks of people from Bangladeshi and Pakistani background similarly in the two waves. Further adjustment for pre-pandemic health status also attenuated the relationship. However, even after full adjustment, people from Pakistani and Bangladeshi background remained substantially at greater risk of COVID-19 deaths than White British people, with HRs of 2.67 [2.36 – 3.02] and 1.99 [1.67 - 2.38]in men and women from Pakistani background, and 2.55 [2.06 - 3.15] and 2.16 [1.60 – 2.91] in men and women from Bangladeshi background, respectively. The adjustments had little impact on the HRs for people from Indian background.

## Discussion

### Summary of findings

In this analysis of 28.9 million adults living in private households and 46,790 COVID-19 related deaths, we highlight several major findings. First, in the first wave all ethnic minority groups were at elevated risk of COVID-19 related death, and in the second wave, people from South Asian background, in particular Bangladeshi and Pakistani, but not Black individuals, were at greater risk of COVID-19 death compared to the White British population. Second, geographical factors explained more than half of the differences in COVID-19 mortality risk in the first wave, but much less in the second wave. Third, socio-demographic factors explained a similar proportion of the elevated risks of people from Bangladeshi and Pakistani background in the first and second waves. Fourth, adjusting for comorbidities did not substantially reduce the ethnic difference in risk of COVID-19 related death, after other factors that had already been accounted for.

### Comparison with related studies

In line with existing studies investigating ethnic inequalities in SARS-CoV-2 infection and COVID-19 mortality [15, 3, 4, 16, 17], we find that most ethnic minority groups were disproportionally affected in the first wave. Our findings that the ethnic inequalities in COVID-19 mortality differed between the two waves is consistent with the evidence that these disparities are likely to be driven by differences in exposure to infection and therefore can change over time. Existing evidence suggests that the lockdown measures implemented in March 2020 were associated with a reduction in inequalities in mortality in England in all ethnic minority groups [3].

Several studies analysed the ethnic inequalities in COVID-19 mortality in the first wave, adjusting for detailed socio-demographic factors [3] or detailed pre-existing health conditions [4]. Our study is the first to investigate simultaneously the role of socio-demographic factors and health conditions in explaining the differences in COVID-19 mortality between ethnic groups between the first and the second wave in a large nationwide population. We find that after adjusting for geographical and socio-demographic factors, adjusting for pre-existing conditions only moderately reduced the estimated differences in COVID-19 mortality between ethnic groups. This suggests that these inequalities in mortality are primarily driven by differences in exposure and infection, which is corroborated by findings from a study based on antibody testing [17].

### Strengths and limitations

The primary strength of our study is the use of a unique, nationwide, newly linked population-level data set based on the General Practice Extraction Service (GPES) Data for pandemic planning and research, linked to the most comprehensive and reliable sources of sociodemographic variables from the latest census, mortality records and Hospital Episode Statistics. Unlike studies based solely on electronic health records, our study is based on self-identified ethnicity, with very few missing data. Our data contain both detailed socio-demographic characteristics, such as household composition, housing quality, and occupational exposure, and extensive information on pre-pandemic health based on primary care and hospital records. To our knowledge, our study is the first to use nationally representative linked data to examine the association between ethnicity and COVID-19 mortality while accounting for the effect of both socio-demographic factors and comorbidities.

The main limitation of our study data set is the 9-year lag between census day and the start of the pandemic. Most socio-demographic characteristics included in our models reflect the situations of individuals as they were in 2011, not necessarily those at the start of the COVID-19 pandemic. To mitigate this, we excluded people aged less than 30 years old, whose circumstances are the most likely to have changed since the Census. We also updated place of residence based on information from the 2019 NHS Patient Register. Since the socio-demographic factors are less likely to have changed for older people than younger people, measurement error is likely to be smaller for the people at greater risk. Another limitation is that the study population is limited to people enumerated at the 2011 Census, and therefore did not include people who immigrated or were born between 2011 and 2020. As a result, it did not fully represent the population at risk. However, migrants tend to be young and the risk of COVID-19 mortality is low for young people [12].

### Mechanisms

We find that in the second wave the disparities are more pronounced in people of South Asian ethnicity particularly those from Pakistani and Bangladeshi backgrounds. Compared to people from other ethnic groups, these groups are more likely to reside in deprived areas, in large households and in multigenerational families [3]. Households are important contributor to transmission of COVID-19, with household size being associated with risk of SARS-CoV-2 infection [18, 19, 20]. Secondary attack rates within household are high [21], and as a result living in multi-generational household is associated with increased risk of COVID-19 mortality amongst elderly adults in England [22]. Differences in occupational exposure could also account for some of the differences in mortality between groups, as a higher proportion of Pakistani and Bangladeshi men work as taxi drivers, shopkeepers and proprietors than any other ethnic backgrounds [23]. Previous research showed that ethnic minority groups also experience other structural factors that increase their likelihood of risk of mortality. [24].

Whilst our study adjusts for a range of socio-demographic factors, including household composition and occupational exposure, we may not capture fully the effect of these factors because of measurement error. Our study also accounts for differences in pre-pandemic health. Potential contributing factors not measured in our data include linguistic and cultural factors as well as barriers to accessing public health messaging [25]. Further research, including qualitative studies, would be needed to understand better the differences observed between the waves.

### Implications of the findings

The finding of a strong reduction in the difference in COVID-19 mortality between people from Black ethnic background and people from the White British group is reassuring. The widespread dissemination of research findings and government reports published during the first wave of infection that highlighted that people form ethnic minority groups were disproportionally affected by COVID-19 may have helped raise the awareness of these disparities amongst the general public. However, the continued higher rate of mortality in people from Bangladeshi and Pakistani background is alarming, and requires focused public health campaign and policy response. Focusing on treating underlying conditions, although important, may not be enough to reduce the inequalities in COVID-19 mortality. Understanding the need of these ethnic groups, through engagement with local communities, public health and healthcare teams, must be at the core of any public health response.

## Conclusion

Our study showed that the risk of COVID-19 mortality during the first wave of COVID-19 pandemic was higher in people from ethnic minority background, both in men and women, compared to people from White ethnic background. There was a reduction of COVID-19 mortality during the second wave in most of the ethnic groups while the higher rates continued in men and women from Bangladeshi and Pakistani background. Focused public health policy may help reduce the existing and widening inequalities in COVID-19 mortality.

## Data Availability

The ONS Public Health Linked Data Asset will be made available on the ONS Secure Research Service for Accredited researchers. Researchers can apply for accreditation through the Research Accreditation Service.

## Footnotes

### Ethics approval

Ethical approval was obtained from the National Statistician’s Data Ethics Advisory Committee (NSDEC(20)12)

### Author contributors

All authors contributed to the study conceptualisation and design. VN lead the preparation of the study data and performed the statistical analyses. All authors contributed to interpretation of the results. VN and NI drafted the manuscript. All authors contributed to the critical revision of the manuscript. All authors approved the final manuscript. VN is the guarantor for the study. The corresponding author attests that all listed authors meet authorship criteria and that no others meeting the criteria have been omitted.

## Supplementary Tables

**Table A1.**
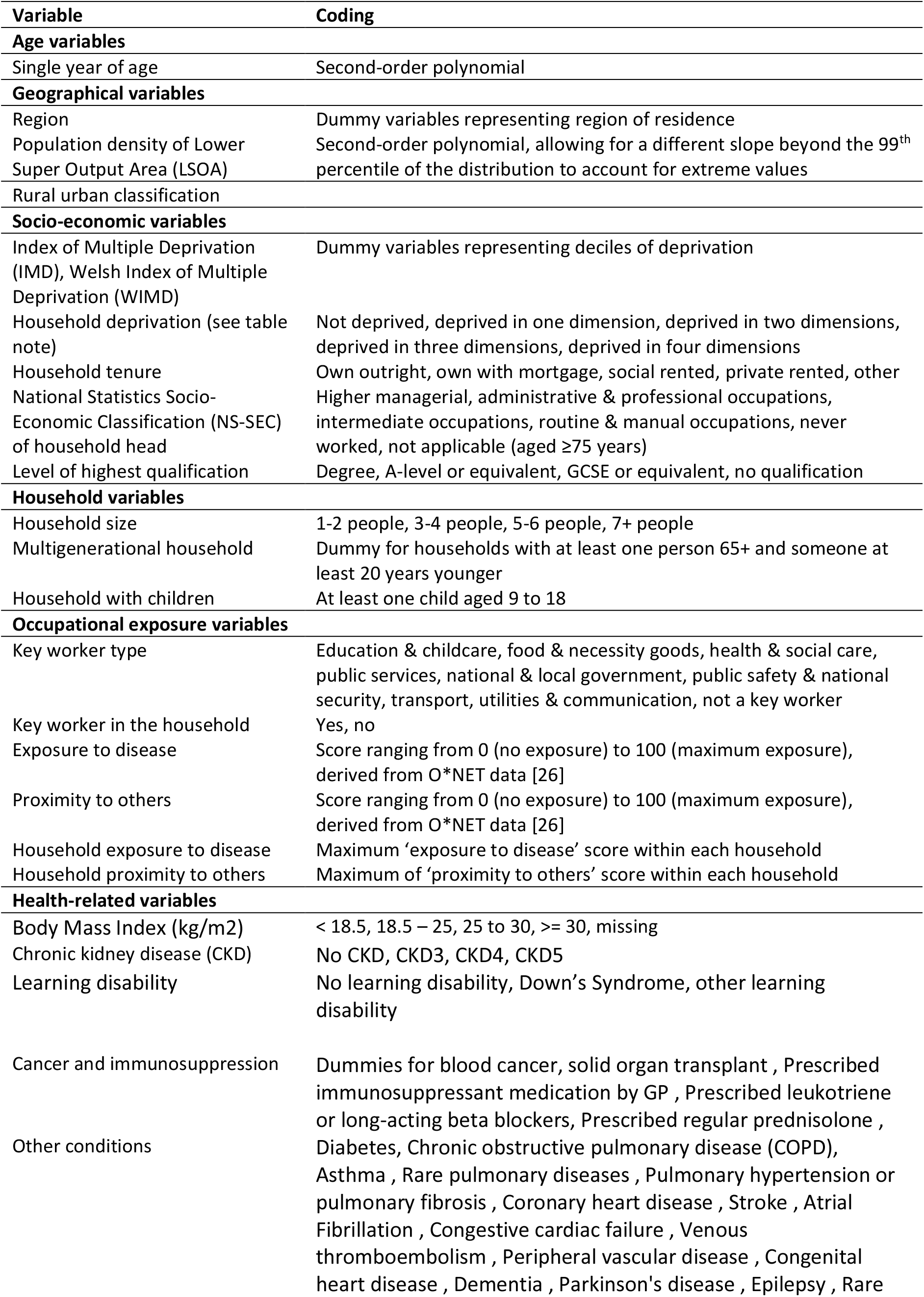
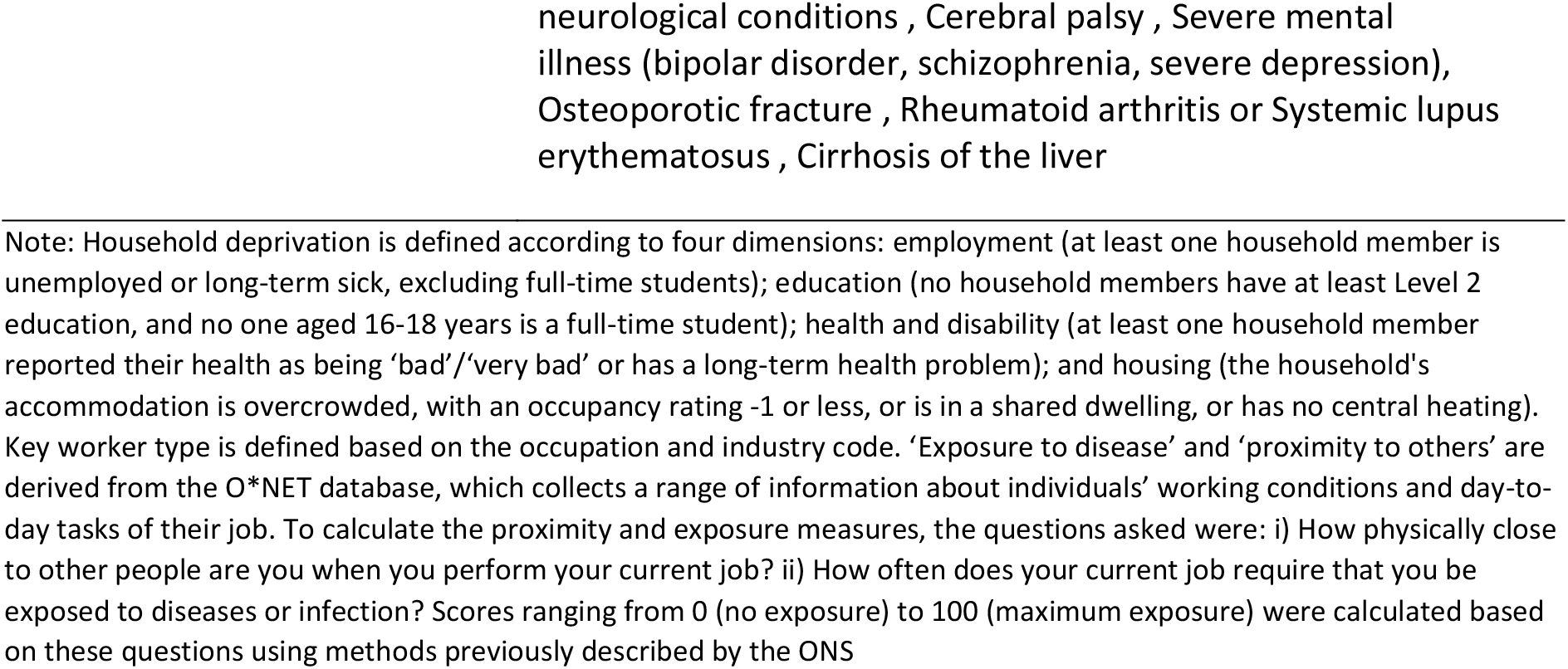
Covariates included in the Cox-regression models

**Table A2.** Mean age at death by ethnic group in the two waves

|  | Wave 1 | Wave 2 |
| --- | --- | --- |
| Bangladeshi | 72.5 | 72.9 |
| Black African | 66.0 | 65.9 |
| Black Caribbean | 77.5 | 77.4 |
| Chinese | 75.3 | 76.4 |
| Indian | 75.1 | 75.4 |
| Mixed | 71.2 | 71.7 |
| Other | 72.4 | 73.1 |
| Pakistani | 72.6 | 73.1 |
| White British | 78.7 | 78.8 |
| White other | 77.8 | 77.8 |

Note: Linked 2011 Census to HES, GPES and Mortality registration data. Sample restricted to individuals 30-100 years old living in private households

**Table A3.**
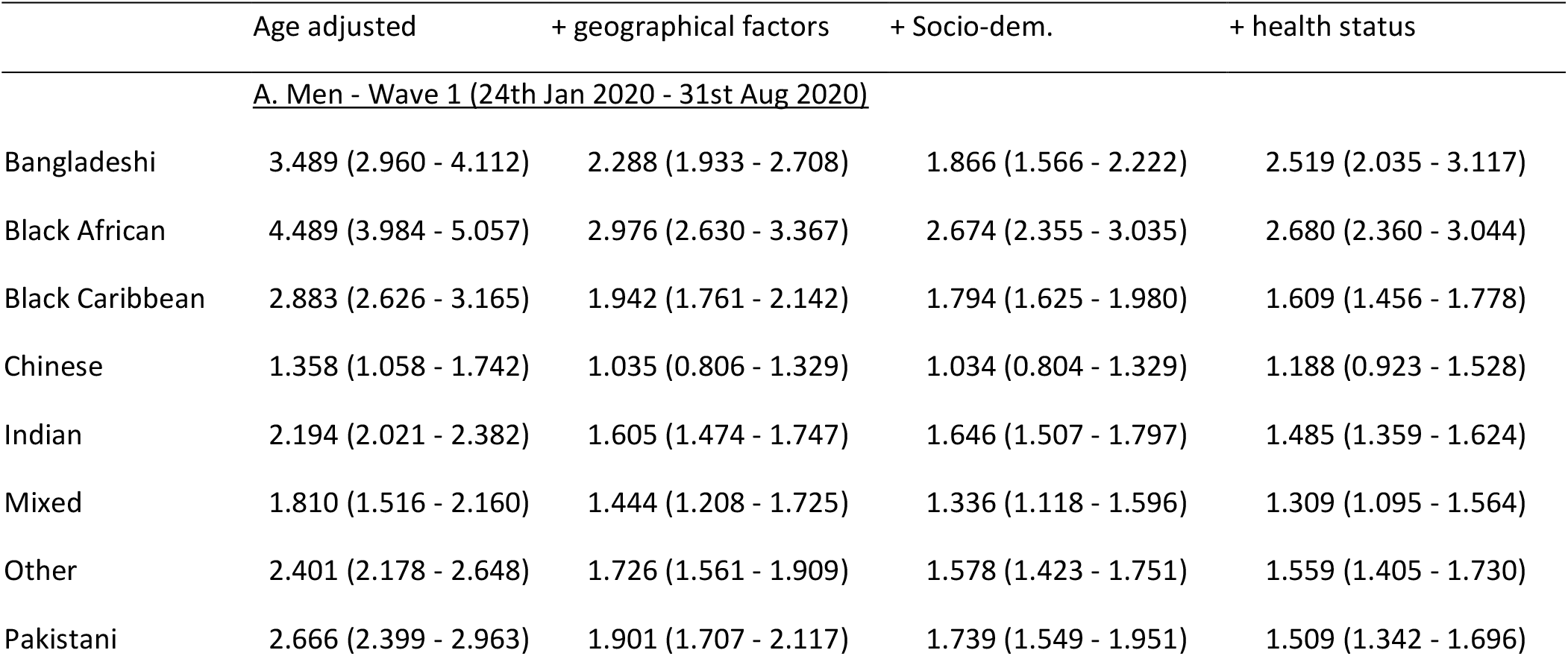
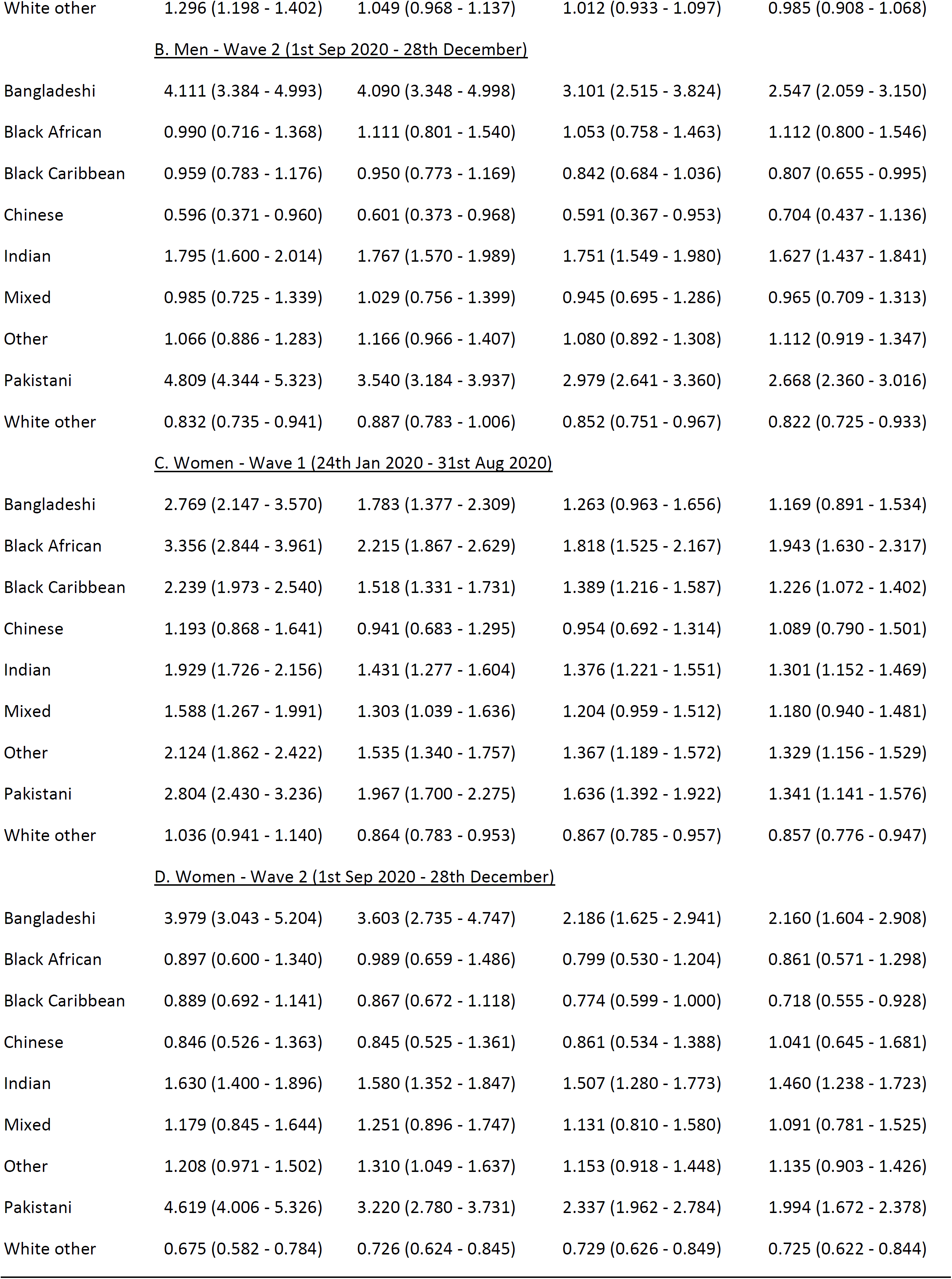
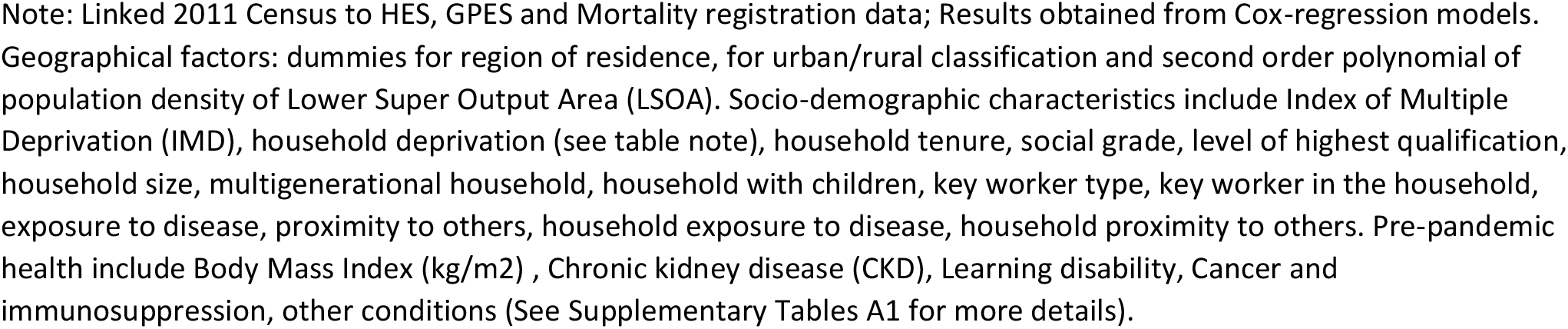
Hazard ratios for COVID-19 related death for ethnic-minority groups compared with the White British population stratified by sex and period

